# The psychological impact of ‘mild lockdown’ in Japan during the COVID-19 pandemic: a nationwide survey under a declared state of emergency

**DOI:** 10.1101/2020.07.17.20156125

**Authors:** Tetsuya Yamamoto, Chigusa Uchiumi, Naho Suzuki, Junichiro Yoshimoto, Eric Murillo-Rodriguez

## Abstract

This study examined the psychological distress caused by non-coercive lockdown (mild lockdown) in Japan. An online survey was conducted with 11,333 people (52.4% women; mean age = 46.3 ± 14.6 years, range = 18-89 years) during the mild lockdown in the seven prefectures most affected by COVID-19 infection. Over one-third (36.6%) of participants experienced mild-to-moderate psychological distress (Kessler Psychological Distress Scale [K6] score 5-12), while 11.5% reported serious psychological distress (K6 score ≥ 13). The estimated prevalence of depression (Patient Health Questionnaire-9 score ≥ 10) was 17.9%. Regarding the distribution of K6 scores, the proportion of individuals displaying psychological distress in this study was significantly higher compared to previous national survey data from 2010, 2013, 2016 and 2019. Healthcare workers, those with a history of treatment for mental illness, and younger participants (aged 18-19 or 20-39 years) were particularly vulnerable. Psychological distress severity was influenced by specific interactional structures of risk factors: high loneliness, poor interpersonal relationships, COVID-19-related sleeplessness and anxiety, deterioration of household economy, and work and academic difficulties. Flexible approaches that are optimised for the difficulties specific to each individual through cross-disciplinary public-private initiatives are important to combat lockdown-induced mental health problems.

## Introduction

Given the spread of the coronavirus disease 2019 (COVID-19) infection, as of June 2020, the number of infected people worldwide is still increasing ^1^. Although outbreaks have subsided in some areas of Europe and East Asia, the threat of a new wave of infections remains a serious concern. Therefore, there is an urgent need to accumulate research on the effects of lockdowns (urban blockades), which should be used as a reference in policymaking during the spread of infection. While the lockdowns that have been implemented so far have been effective in limiting the spread of infection, many negative psychological effects of lockdowns (e.g., stress, loneliness) exist ^2–6^ and there is room for improvement in lockdown implementation.

Under these circumstances, it may be useful to examine the impact of a ‘mildly enforceable lockdown’ in Japan. A total of 4,111 infections and 97 deaths were confirmed in Japan by 6 April 2020 ^7^. On 7 April 2020, the Japanese government declared a state of emergency for the first time. This authorises prefectural governors to ‘request’ (or ‘instruct’ if residents do not comply) residents to (1) refrain from going out of their homes for non-essential reasons and (2) restrict the use of stores and facilities. Enforceable measures are extremely limited in Japan’s emergency declarations and are much less restrictive than the ‘lockdowns’ introduced in some areas of Europe and the United States. There are no penalties for disobedience. Therefore, citizens are obliged to try to cooperate with measures taken by prefectures, such as voluntarily taking time off work and refraining from going out. Here, we defined ‘mild lockdown’ as a state of lockdown specific to Japan relying on voluntary public cooperation.

There is a high prevalence of psychological symptoms such as depression and anxiety among people who experienced lockdown during the COVID-19 pandemic ^2–6^, and containment measures against such a pandemic can have a strong impact on individuals’ daily lives and their psychological well-being ^8^. However, in studies examining the psychological effects of lockdowns reported to date, lockdowns have been accompanied by coercive forces, and the effects of mild lockdown remain unclear. With the current state of alert for further spread and the potential for a second wave of COVID-19, it is vital to clarify the effects of mild lockdown on people’s mental health to consider future prevention policies and appropriate intervention strategies. Therefore, this study identified psychological distress severity and its risk and protective factors during mild lockdown.

The timing of data collection and the selection of target areas are important in examining the effects of mild lockdown, which change daily. We selected the data collection as the period from the start of mild lockdown—based on the declaration of a state of emergency—until mild lockdown phasing out began (i.e. 7 April 2020 to 12 May 2020). After our data collection was completed, the mild lockdown was phased out on 14 May 2020, and it was fully lifted on 25 May 2020. Therefore, our data collection period was in the middle of the mild lockdown—a period of great distress and less susceptible to recall bias. Furthermore, due to regional differences in the spread of infection, we included residents from the seven prefectures initially subject to the declaration of a state of emergency (Tokyo, Kanagawa, Osaka, Saitama, Chiba, Hyogo, and Fukuoka), all in heavily populated and heavily affected areas. We examined the impact of mild lockdown on the population by identifying the distribution of psychological distress severity in the target areas during these periods and comparing it with data previously collected by the government.

There is also an urgent need to identify the impact on those considered vulnerable (e.g. healthcare workers and older people) to determine appropriate responses to the difficulties faced by vulnerable populations during these pandemics ^8,9^. While previous studies report higher negative mental health risks among healthcare workers ^8,10,11^, there remain inconsistencies for the psychological impact on young and older adults ^6,12,13^; thus, more research is needed. We therefore, examined psychological distress caused by mild lockdown, focusing on healthcare workers, family members of healthcare workers, those undergoing treatment for, or with a history of, physical or mental illness, and older adults (aged ≥ 65) and younger adults (18-19) who have previously been identified as potentially vulnerable ^9^.

It is important to consider psychosocial variables, such as stressors and stress management strategies specific to lockdown, in identifying factors that influence the impact of mild lockdown. Given that such variables have not previously been adequately considered ^3,5,6^ we examined the risk and protective factors for psychological distress, including psychosocial variables such as life changes due to lockdown and lifestyle habits during lockdown.

Additionally, we assumed that various risk and protective factors are intricately related and that people may have diverse backgrounds of psychological distress in lockdown situations. It is important to understand the background of psychological distress to consider approaches tailored to individuals’ difficulties. However, no previous studies have elaborated on this. Therefore, we used non-parametric Bayesian co-clustering ^14^—a method of unsupervised learning. This method allows exhaustive visualisation of the underlying interaction structure among many variables. Therefore, it was expected to elucidate various problem structures that cause psychological distress during mild lockdown.

In sum, given that there are currently no research findings specifically addressing the impact of mild lockdown, this study is useful in that it clarifies the impact of mild lockdown on various populations and provides basic data that will be useful in formulating optimal strategies during future periods of infection spread and pandemics.

## Methods

### Participants and data collection

A total of 11,333 participants (mean age = 46.3±14.6 years, range = 18-89) were included for analyses. Participants’ socio-demographic characteristics are described in Table 1. The survey was conducted online between 11 May and 12 May 2020 and was designed to assess the psychological impact of the mild lockdown on participants over approximately one month—from the start of the mild lockdown (7 April 2020) to its phasing out in some areas (14 May 2020). Through Macromill.inc., approximately 80,000 people were recruited by email, and data were collected on an online platform. To sensitively detect the impact of mild lockdown, participants were recruited only in the seven prefectures where the emergency declaration was first applied (Tokyo, Kanagawa, Osaka, Saitama, Chiba, Hyogo, and Fukuoka). These cities were assumed to be susceptible to mild lockdown given their large populations and the large number of cases reported in these cities.

**Table 1.** Participants’ socio-demographic characteristics.

| Characteristics, *n* (%) | Total | Psychological distress | | | $\chi^2$ | df | *p* |
| --- | --- | --- | --- | --- | --- | --- | --- |
|  |  | No or low | Mild-to-moderate | Serious |  |  |  |
| **Overall** | 11333 | 5884 (51.9) | 4146 (36.6) | 1303 (11.5) | 2831.00 | 2 | <0.001 |
| **Age** |  |  |  |  |  |  |  |
| 18-19 | 143 | 62 (43.4) | 50 (35.0) | 31 (21.7) | 428.66 | 6 | <0.001 |
| 20-39 | 3745 | 1633 (43.6) | 1508 (40.3) | 604 (16.1) |  |  |  |
| 40-64 | 6024 | 3157 (52.4) | 2230 (37.0) | 637 (10.6) |  |  |  |
| ≥ 65 | 1421 | 1032 (72.6) | 358 (25.2) | 31 (2.2) |  |  |  |
| **Sex** |  |  |  |  |  |  |  |
| Male | 5391 | 3098 (57.5) | 1789 (33.2) | 504 (9.3) | 134.68 | 2 | <0.001 |
| Female | 5942 | 2786 (46.9) | 2357 (39.7) | 799 (13.4) |  |  |  |
| **Occupation category** |  |  |  |  |  |  |  |
| Employed | 7685 | 3948 (51.4) | 2852 (37.1) | 885 (11.5) | 75.60 | 8 | <0.001 |
| Homemaker | 1806 | 919 (50.9) | 699 (38.7) | 188 (10.4) |  |  |  |
| Student | 407 | 175 (43.0) | 158 (38.8) | 74 (18.2) |  |  |  |
| Unemployed | 1068 | 662 (62.0) | 304 (28.5) | 102 (9.6) |  |  |  |
| Other | 367 | 180 (49.0) | 133 (36.2) | 54 (14.7) |  |  |  |
| **Healthcare worker(self)** |  |  |  |  |  |  |  |
| Yes | 661 | 299 (45.2) | 288 (43.6) | 74 (11.2) | 15.46 | 2 | <0.001 |
| No | 10672 | 5585 (52.3) | 3858 (36.2) | 1229 (11.5) |  |  |  |
| **Healthcare worker(family)** |  |  |  |  |  |  |  |
| Yes | 991 | 493 (49.7) | 373 (37.6) | 125 (12.6) | 2.49 | 2 | 0.2874 |
| No | 10342 | 5391 (52.1) | 3773 (36.5) | 1178 (11.4) |  |  |  |
| **Marital status** |  |  |  |  |  |  |  |
| Married | 7043 | 3933 (55.8) | 2500 (35.5) | 610 (8.7) | 191.36 | 2 | <0.001 |
| Unmarried | 4290 | 1951 (45.5) | 1646 (38.4) | 693 (16.2) |  |  |  |
| **Annual household income (JPY)** |  |  |  |  |  |  |  |
| < 2.0 million | 633 | 273 (43.1) | 233 (36.8) | 127 (20.1) | 149.24 | 10 | <0.001 |
| 2.0-3.9 million | 1990 | 1020 (51.3) | 739 (37.1) | 231 (11.6) |  |  |  |
| 4.0-5.9 million | 2214 | 1174 (53.0) | 797 (36.0) | 243 (11.0) |  |  |  |
| 6.0-7.9 millin | 1495 | 817 (54.6) | 529 (35.4) | 149 (10.0) |  |  |  |
| ≥ 8.0 million | 2130 | 1267 (59.5) | 694 (32.6) | 169 (7.9) |  |  |  |
| Unknown | 2871 | 1333 (46.4) | 1154 (40.2) | 384 (13.4) |  |  |  |
| **Current treatment of severe physical diseases** |  |  |  |  |  |  |  |
| Yes | 482 | 248 (51.5) | 168 (34.9) | 66 (13.7) | 1.85 | 2 | 0.3972 |
| No | 10851 | 5636 (51.9) | 3978 (36.7) | 1237 (11.4) |  |  |  |
| **Previous treatment of severe physical diseases** |  |  |  |  |  |  |  |
| Yes | 851 | 440 (51.7) | 304 (35.7) | 107 (12.6) | 1.12 | 2 | 0.5709 |
| No | 10482 | 5444 (51.9) | 3842 (36.7) | 1196 (11.4) |  |  |  |
| **Current treatment of psychological problems** |  |  |  |  |  |  |  |
| Yes | 641 | 110 (17.2) | 271 (42.3) | 260 (40.6) | 663.31 | 2 | <0.001 |
| No | 10692 | 5774 (54.0) | 3875 (36.2) | 1043 (9.8) |  |  |  |
| **Previous treatment of psychological problems** |  |  |  |  |  |  |  |
| Yes | 1366 | 383 (28.0) | 600 (43.9) | 383 (28.0) | 563.09 | 2 | <0.001 |
| No | 9967 | 5501 (55.2) | 3546 (35.6) | 920 (9.2) |  |  |  |

The number of people collected in each prefecture was determined according to the ratio of the number of people living in each: Tokyo (n = 2,783, 24.6%), Kanagawa (n = 1,863, 16.4%), Osaka (n = 1,794, 15.8%), Saitama (n = 1,484, 13.1%), Chiba (n = 1,263, 11.1%), Hyogo (n = 1,119, 9.9%), and Fukuoka (n = 1,027, 9.1%). The exclusion criteria for participants were; aged <18 years; high school students; and living outside the seven prefectures. The online survey was completed on the second day after link distribution. All participants voluntarily responded to the anonymous survey and provided informed consent online. The survey procedure was clearly explained, and participants could interrupt or terminate participation at any time without explanation. This study was approved by the Research Ethics Committee at the Graduate School of Social and Industrial Science and Technology, Tokushima University (no.212) and was performed according to the ethical standards of the 1964 Declaration of Helsinki and its amendments.

We used published data from a previous Comprehensive Survey of Living Conditions (CSLC) ^15^ to examine changes in psychological distress severity due to mild lockdown. The CSLC is a national survey conducted by the Ministry of Health, Labour and Welfare to assess the health status of the Japanese population. In the CSLC, the Kessler Psychological Distress Scale (K6) ^16^ was used to measure psychological distress. Based on their score classification (0-4, 5-9, 10-14, over 15), the percentages of people in that classification for 2010, 2013, 2016 and 2019 are now publicly available ^15^. These results were used to compare to the survey data in this study. The CSLC data were compiled from a sample of 228,864 households in 2010, 234,383 households in 2013, 224,208 households in 2016 and 217,179 households in 2019.

## Measurements

### Psychological distress

Psychological distress was measured by the Japanese version of the K6 ^17^, a six-item screening scale of nonspecific psychological stress in the past 30 days. Each question was rated on a scale of 0 (*none of the time*) to 4 (*all of the time*); total scores range from 0-24. Given its brevity and high accuracy, the K6 is an ideal scale for screening for mental disorders in population-based health surveys ^17–19^. Additionally, because the duration of symptoms examined by this scale (the past 30 days) corresponds to the period between the start of mild lockdown and the implementation of the survey (∼1 month), the scale could sensitively reflect the influence of psychological distress caused by mild lockdown.

We adopted a threshold of five points commonly used to screen for mild-to-moderate mood/anxiety disorders ^20^. K6 scores ranging from 5-12 were defined as mild-to-moderate psychological distress (MMPD). This threshold is the optimal lower threshold cut-point for screening for moderate psychological distress ^20^. MMPD was assessed given the risk of progression to more severe disability as well as current distress and disability ^21^. Additionally, to screen for severe mood/anxiety disorders, we adopted a threshold score of 13, a criterion traditionally used.^18,22^. A score of ≥ 13 was defined as serious psychological distress (SPD). Additionally, a score of <4 was defined as no or low psychological distress (NPD). Based on 3 years of published data concerning K6 from the Ministry of Health, Labour and Welfare^15^, we defined MMPD or SPD (K6≥ 5) as ‘psychological distress’ together, to make comparisons corresponding to the cut-point of K6 severity.

We also used the Japanese version of the Patient Health Questionnaire-9 (PHQ-9) ^23^ to collect other basic information on mental health. The PHQ-9 consists of nine questions, and participants reported depressive symptoms during the past four weeks assessed by a score of 0 (*not at all*) to 3 (*nearly every day*) ^24^. We defined a score of ≥ 10, previously recommended ^23^, as a cut-point, meaning that a person is more likely to have major depression. The PHQ-9 is widely used internationally as a screening scale for depression ^25^ with high reliability and validity ^23^.

### Loneliness and social networks

Loneliness and social networks are key factors associated with mental health ^26–28^ and may affect people’s mental health in mild lockdown ^9,11^. We measured loneliness and social networks using the Japanese version of the UCLA loneliness scale version 3 (UCLA-LS3) ^29^ and the Japanese version of the abbreviated Lubben Social Network Scale (LSNS-6) ^30^, respectively.

The UCLA-LS3 consists of 10 items, each rated from 1 (*never*) to 4 (*always*) ^31^. The scores range from 10-40, with higher scores indicating higher levels of loneliness. The UCLA-LS3 is highly reliable and valid ^29^ and is internationally used for measuring loneliness ^32–34^. The LSNS-6 consists of three items related to family networks and three items related to friendship networks. The number of people in the network is calculated using a six-point scale (0 = *none* to 5 = *nine or more*) for each item ^35^. Scores range from 0-30 points, with higher scores indicating a larger social network and <12 points indicating social isolation. The LSNS-6 is highly reliable and valid^30^ and has been used in many countries ^36–38^.

### Socio-demographic data

Socio-demographic information collected included age, sex, occupation, marital status, and income. To compare the impact on groups assumed vulnerable to the effects of lockdown in previous studies ^6,8,9,11^, information was collected on whether the individual or a family member was a healthcare worker, whether the individual was currently being treated for a psychiatric or physical illness, and whether the individual had a history of previous treatment for psychiatric or physical illness.

### Lifestyle, stress management, and stressors related to mild lockdown

Based on previous literature regarding the COVID-19 pandemic ^3,6,8,11^, we developed eight lifestyle and stress management items (e.g. exercise; ‘I exercised for my health (whether indoors or outdoors)’) and seven stressors (e.g. deterioration of household economy; ‘The family budget has tightened’) that were assumed to be associated with mild lockdown (Table 2). We asked participants to rate the frequency of implementation and experience of these items from the start of the mild lockdown to the time of the survey on a scale of 1 (*not at all*) to 7 (*extremely*).

**Table 2.**
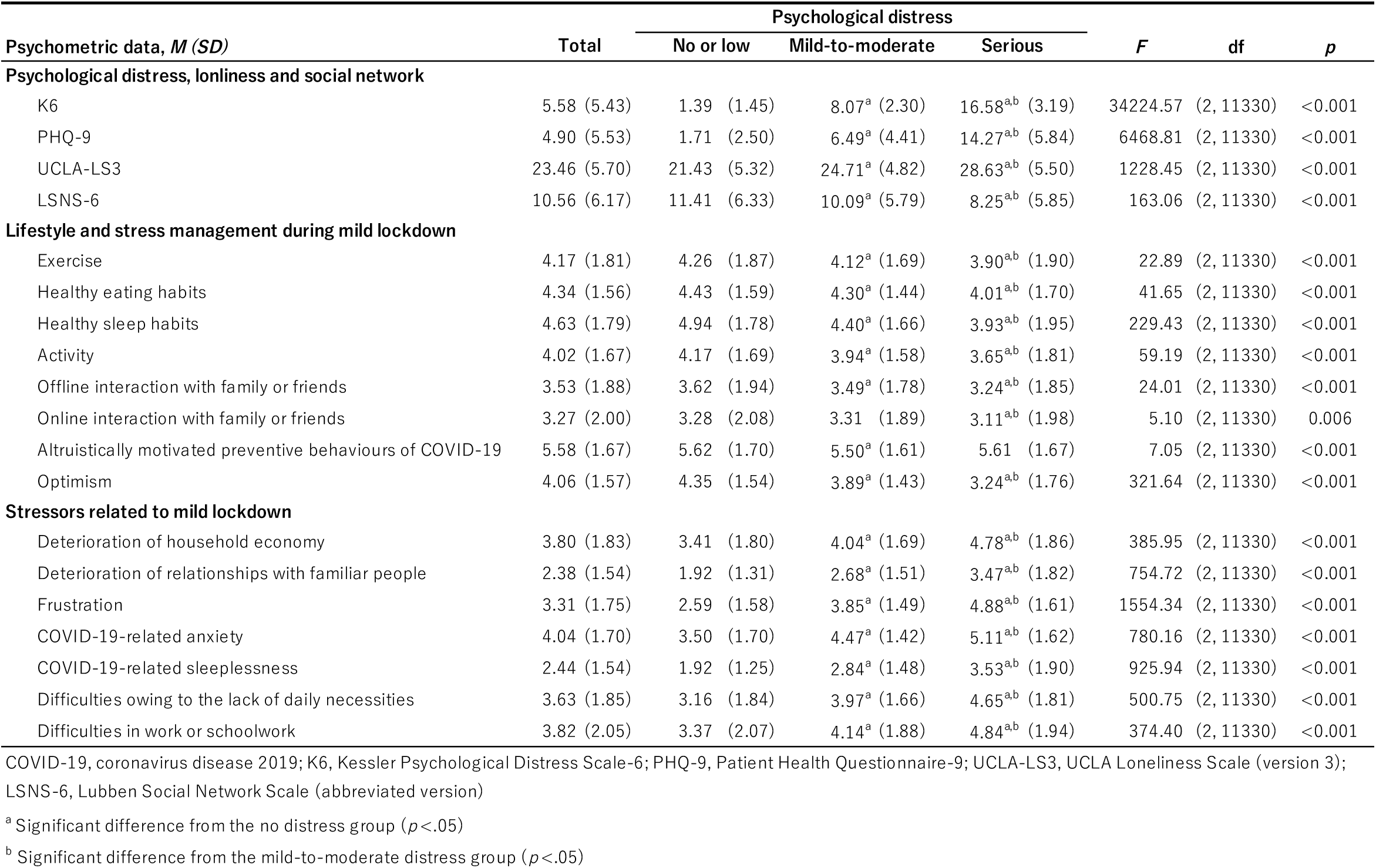
Participants’ psychometric characteristics.

### Statistical analyses

To determine the socio-demographic and psychological characteristics of groups classified by psychological distress severity (NPD (K6 score ≤ 4), MMPD (K6 score 5-12), and SPD (K6 score ≥ 13)), we compared these characteristics using Pearson’s χ^2^ tests for categorical variables and analyses of variance (ANOVAs) for continuous variables. Hochberg’s GT2 method was used for multiple comparisons of ANOVAs to account for differences in numbers between groups. We also used Pearson’s χ^2^ tests against published CSLC data and data from this study to compare the change in the proportion of those with mental illness (K6 ≥ 5) before and during the mild lockdown (2010, 2013, 2016, 2019 and 2020).

To elaborate on psychological distress severity in the group assumed to be vulnerable, we conducted ANOVAs with K6 scores as the dependent variable for each of the following categories: healthcare worker (individual, family, individual and family, and none), psychiatric illness (currently treated, previously treated, both, and none), physical illness (currently treated, previously treated, both, and none), and age in years (18-19, 20-39, 40-64, and ≥ 65). Hochberg’s GT2 method was used for multiple comparisons to account for differences in numbers between groups.

Multinomial logistic regression analyses were conducted to examine the effects on psychological distress of socio-demographic characteristics (age, sex, healthcare workers, and history of treatment for mental and physical illness) that previous studies suggest increased vulnerability, and psychosocial variables related to mild lockdown, including loneliness and social networks. Based on Field ^39^, the model was examined using the forward entry method, and the final model was constructed by employing variables that significantly contribute to the explanation of the model. Multicollinearity among the independent variables of the final model was checked to assess potential bias in the results due to collinearity.

Non-parametric Bayesian co-clustering ^14^ was used to visualise the exhaustive interaction structure between the psychosocial variables that were significant in multinomial logistic regression and psychological distress during mild lockdown. Iterations based on the Bayesian optimisation principle were performed 10,000 times to calculate the log marginal likelihood, which indicates the goodness-of-fit of the model. The log marginal likelihoods were completed among the models, and the model with the highest log marginal likelihood was adopted.

For all tests, significance was set at *α* =0.05 (two-tailed). Analyses and figures were drawn using SPSS version 22.0 (SPSS Japan Inc., Tokyo, Japan), MATLAB R2017a (Mathworks Inc.), and RStudio version 1.1.442 ^40,41^.

## Results

### Socio-demographic characteristics by psychological distress severity

The socio-demographic characteristics by severity of psychological distress, as measured by the K6, are shown in Table 1. In total, 4,146 participants (36.6%) had MMPD (K6 score 5-12) and 1,303 participants (11.5%) had SPD (K6 score ≥ 13). The estimated prevalence of depression (PHQ-9 score ≥ 10) was 2,034 (17.9%).

In the MMPD group, significantly more participants were aged 20-39 years (*p* < 0.001), women (*p* < 0.001), employed (*p* = 0.045), homemakers (*p* = 0.020), healthcare worker (*p* < 0.001), single (*p* = 0.001), currently being treated for psychological problems (*p* = 0.001), and had received treatment for psychological problems in the past (*p* < 0.001) as compared to their counterparts.

In the SPD group, the following characteristics were observed to be significantly more prevalent: aged 18-19 years or 20-39 years, women, students, unmarried, income of less than 2 million yen, currently being treated for psychological problems, and having been treated for psychological problems in the past (all *p*s < 0.001).

### Psychometric characteristics by psychological distress severity

Psychological characteristics by psychological distress severity are shown in Table 2. Psychosocial variables that were significantly greater in the MMPD group than in the NPD group (K6 score ≤ 4) included loneliness (UCLA score), deterioration of household economy, deterioration of relationships with familiar people, frustration, COVID-19-related anxiety, COVID-19-related sleeplessness, difficulties due to a lack of daily necessities, and difficulties in work or schoolwork (all *p*s < 0.001). In contrast, psychosocial variables that were significantly less prevalent in the MMPD group than in the NPD group were social network size (LSNS-6 score), exercise, healthy eating habits, healthy sleep habits, activity, offline interaction with family or friends, altruistically motivated preventive behaviours, and optimism (all *p*s < 0.001). Similar results were observed in the SPD group, with the difference being that there was less online interaction with family or friends (*p* = 0.004), and altruistically motivated preventive behaviours were not different from those in the NPD group.

### Chronological comparison of psychological distress

There was a significant difference in the proportion of those with psychological distress (K6 score ≥ 5) in 2010, 2013, 2016, 2019 and 2020 (*χ*^*2*^(8) = 41.9, *p* < 0.001). A residuals analysis revealed that, in 2020, the percentage of NPD and unknown groups was significantly lower (*p* = 0.006; *p* = 0.002, respectively) and the percentage of psychological distress group was significantly higher (*p* < 0.001; Figure 1). Additionally, in 2010, the percentage of unknown groups was significantly higher (*p* < 0.001).

**Figure 1.**
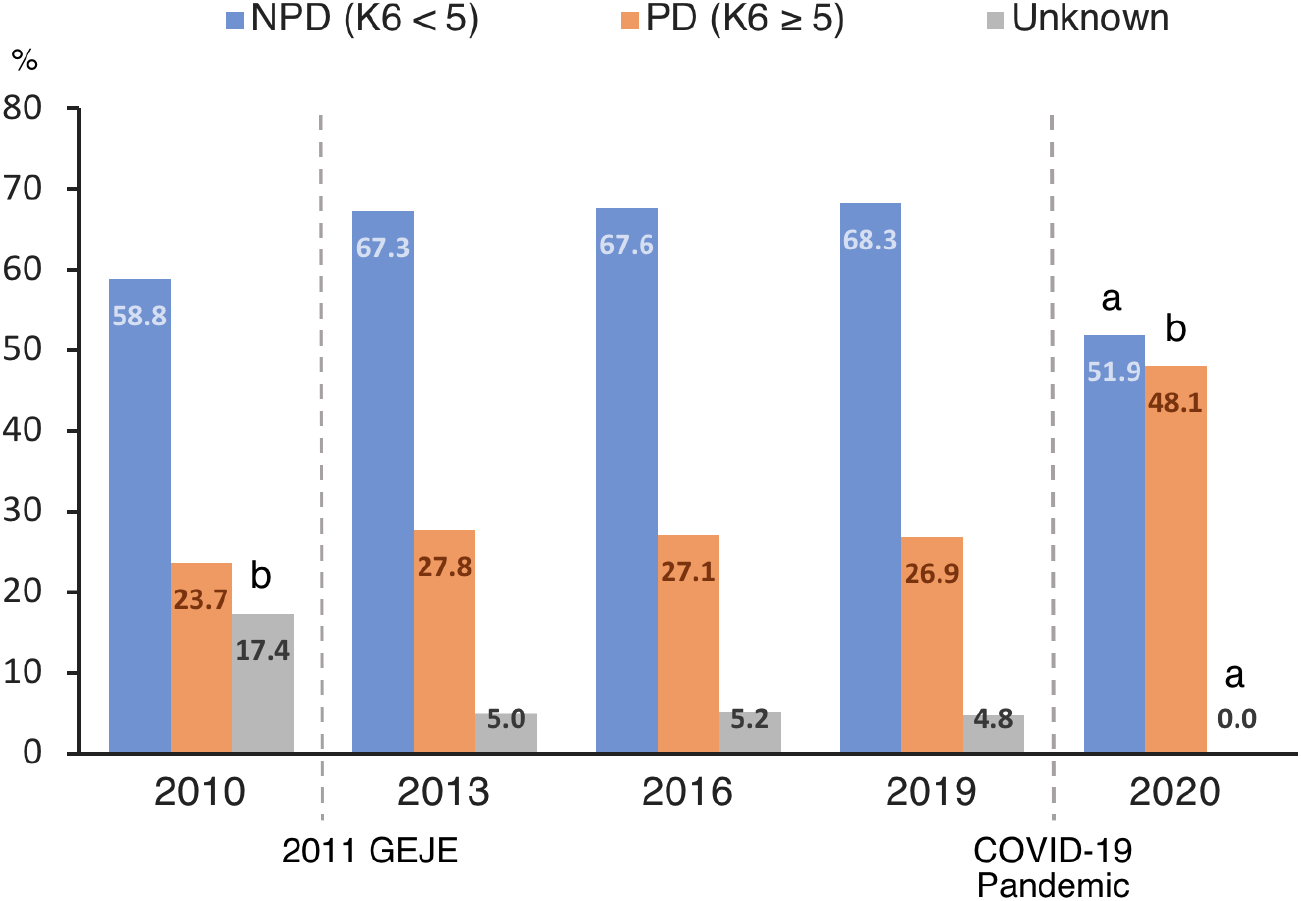
Chronological comparison of the distribution of psychological distress in Japan. COVID-19, coronavirus disease 2019; GEJE, the Great East Japan Earthquake; NPD, no or low psychological distress; PD, psychological distress ^a^ Significantly smaller percentage (*p* < .05) ; ^b^ Significantly larger percentage (*p* < .05).

### Psychological distress in vulnerable groups

The distribution of psychological distress (K6 score) in each group is shown in Figure 2. A one-way ANOVA revealed a significant difference between groups related to the healthcare workers (*F*(3,11,329)= 3.50, *p =* 0.015, η^2^_p_ = 0.001) and those who were healthcare workers had significantly more psychological distress than those who were not healthcare workers (*p =* 0.049; Figure 2A).

**Figure 2.**
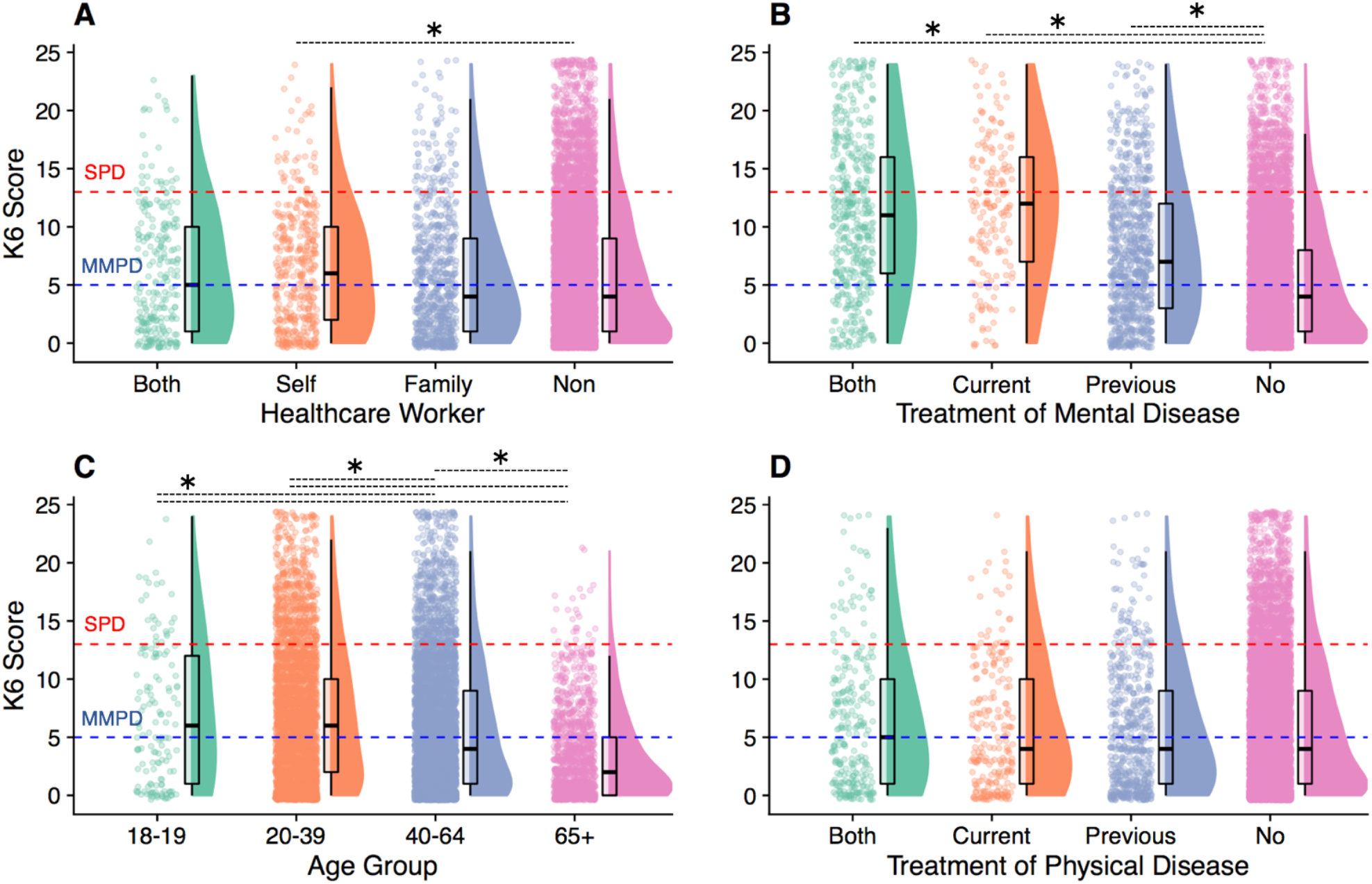
The distribution of psychological distress in the vulnerable groups. MMPD, mild-to-moderate psychological distress; SPD, serious psychological distress The red dotted line indicates the cut-point for SPD (K6 score ≥ 13) and the blue dotted line indicates the cut-point for MMPD (K6 score 5-12). * *p* < 0.05, two-tailed.

There was also a significant difference between the groups related to the treatment of psychiatric disorders (*F*(3, 11,329) = 359 .32, *p* < 0.001, η^2^_p_ = 0.087). Significantly more psychological distress was reported by those who were currently being treated for psychological problems, had ever been treated for psychological problems, or both, than by those who had not been treated for psychological problems (all *p*s < 0.001; Figure 2B).

There was also a significant difference among age group (*F*(3, 11,329) = 159.22, *p* < 0.001, η^2^_p_ = 0.040): psychological distress was higher for those aged 18-19, 20-39, and 40-64 years than those aged ≥ 65 (all *p*s < 0.001; Figure 2C). Psychological distress was also higher in those aged 18-19 and 20-39 than those aged 40-64 (*p*s = 0.010 and < 0.001, respectively; Figure 2C). There was no significant between-group difference in psychological distress concerning the treatment of physical diseases (*F*(3,11,329) = 1.51, *p =* 0.210, η^2^_p_= 0.000; Figure 2D).

### Risk and preventive factors for psychological distress

The results of the final multinomial logistic regression model are shown in Table 3. No multicollinearity problems were found among the independent variables (all variance inflation factors < 1.87).

**Table 3.** Multinominal logistic regression analysis between psychological distress and related factors.

| Predictor | $\beta$ (SE) | OR [95% CI] | p |
| --- | --- | --- | --- |
| **Mild to Moderate Psychological Distress** |  |  |  |
| **Age** |  |  |  |
| >65 (ref.) | 0 |  |  |
| 18-19 | 0.34 (0.23) | 1.40 [0.89 - 2.20] | 0.141 |
| 20-39 | 0.31 (0.09) | 1.36 [1.14 - 1.62] | 0.001 |
| 40-64 | 0.11 (0.08) | 1.11 [0.95 - 1.30] | 0.179 |
| **Sex** |  |  |  |
| Male (ref.) | 0 |  |  |
| Female | 0.23 (0.05) | 1.25 [1.13 - 1.39] | <0.001 |
| **Healthcare worker (self)** |  |  |  |
| No (ref.) | 0 |  |  |
| Yes | 0.27 (0.10) | 1.31 [1.07 - 1.59] | 0.009 |
| **Previous treatment of severe physical diseases** |  |  |  |
| No (ref.) | 0 |  |  |
| Yes | 0.24 (0.10) | 1.27 [1.06 - 1.53] | 0.012 |
| **Current treatment of psychological problems** |  |  |  |
| No (ref.) | 0 |  |  |
| Yes | 0.95 (0.14) | 2.59 [1.96 - 3.41] | <0.001 |
| **Previous treatment of psychological problems** |  |  |  |
| No (ref.) | 0 |  |  |
| Yes | 0.57 (0.09) | 1.77 [1.50 - 2.10] | <0.001 |
| **Psychosocial variables** |  |  |  |
| UCLA-LS3 | 0.12 (0.01) | 1.13 [1.12 - 1.14] | <0.001 |
| LSNS-6 | 0.02 (0.01) | 1.02 [1.01 - 1.03] | <0.001 |
| Online interaction with family or friends | 0.03 (0.01) | 1.03 [1.01 - 1.06] | 0.017 |
| Optimism | -0.12 (0.02) | 0.89 [0.86 - 0.92] | <0.001 |
| Healthy sleep habits | -0.08 (0.02) | 0.92 [0.90 - 0.95] | <0.001 |
| Deterioration of household economy | 0.04 (0.02) | 1.04 [1.01 - 1.07] | 0.018 |
| Deterioration of relationships with familiar people | 0.06 (0.02) | 1.07 [1.03 - 1.11] | 0.001 |
| Frustration | 0.26 (0.02) | 1.30 [1.26 - 1.35] | <0.001 |
| COVID-19-related anxiety | 0.23 (0.02) | 1.26 [1.22 - 1.30] | <0.001 |
| COVID-19-related sleeplessness | 0.20 (0.02) | 1.22 [1.17 - 1.27] | <0.001 |
| Difficulties in work or schoolwork | 0.04 (0.01) | 1.04 [1.01 - 1.07] | 0.007 |

**Table 3. Cont.**
|  |  |  |  |
| --- | --- | --- | --- |
| **Serious Psychological Distress** |  |  |  |
| **Age** |  |  |  |
| >65 (ref.) | 0 |  |  |
| <20 | 1.91 (0.36) | 6.78 [3.33 - 13.82] | <0.001 |
| 20-39 | 1.25 (0.22) | 3.50 [2.29 - 5.36] | <0.001 |
| 40-64 | 0.58 (0.21) | 1.79 [1.18 - 2.70] | 0.006 |
| **Sex** |  |  |  |
| Male (ref.) | 0 |  |  |
| Female | 0.28 (0.09) | 1.32 [1.11 - 1.56] | 0.002 |
| **Healthcare worker (self)** |  |  |  |
| No (ref.) | 0 |  |  |
| Yes | -0.07 (0.17) | 0.94 [0.67 - 1.32] | 0.707 |
| **Previous treatment of severe physical diseases** |  |  |  |
| No (ref.) | 0 |  |  |
| Yes | 0.36 (0.16) | 1.43 [1.04 - 1.95] | 0.026 |
| **Current treatment of psychological problems** |  |  |  |
| No (ref.) | 0 |  |  |
| Yes | 1.88 (0.17) | 6.58 [4.68 - 9.23] | <0.001 |
| **Previous treatment of psychological problems** |  |  |  |
| No (ref.) | 0 |  |  |
| Yes | 0.98 (0.12) | 2.66 [2.10 - 3.37] | <0.001 |
| **Psychosocial variables** |  |  |  |
| UCLA-LS3 | 0.27 (0.01) | 1.30 [1.28 - 1.33] | <0.001 |
| LSNS-6 | 0.03 (0.01) | 1.03 [1.02 - 1.05] | <0.001 |
| Online interaction with family or friends | 0.08 (0.02) | 1.08 [1.03 - 1.13] | 0.002 |
| Optimism | -0.28 (0.03) | 0.76 [0.71 - 0.80] | <0.001 |
| Healthy sleep habits | -0.14 (0.03) | 0.87 [0.83 - 0.91] | <0.001 |
| Deterioration of household economy | 0.13 (0.03) | 1.14 [1.08 - 1.20] | <0.001 |
| Deterioration of relationships with familiar people | 0.19 (0.03) | 1.21 [1.15 - 1.28] | <0.001 |
| Frustration | 0.53 (0.03) | 1.70 [1.60 - 1.81] | <0.001 |
| COVID-19-related anxiety | 0.40 (0.03) | 1.49 [1.40 - 1.60] | <0.001 |
| COVID-19-related sleeplessness | 0.33 (0.03) | 1.40 [1.32 - 1.48] | <0.001 |
| Difficulties in work or schoolwork | 0.10 (0.03) | 1.11 [1.05 - 1.16] | <0.001 |

Note. $R^2 = 0.41$ (Cox-Snell), 0.48 (Nagelkerke). Model $\chi^2(38) = 5902.04$ , $p < 0.001$
COVID-19, coronavirus disease 2019; UCLA-LS3, UCLA Loneliness Scale (version 3); LSNS-6, Lubben Social Network Scale (abbreviated version)

First, the prominent risk factors (all odds ratios [ORs] > 1.30) that predicted MMPD included being aged 20-39, a healthcare worker and treatment for psychological problems either currently or in the past. Protective factors were optimism (OR = 0.89, 95% confidence interval [CI] = 0.86-0.92, *p* < 0.001) and healthy sleep habits (OR = 0.92, 95% CI = 0.90-0.95, *p* < 0.001).

Next, the prominent risk factors (all ORs ≥ 1.30) that predicted SPD were indicated as follows: aged 18-19, 20-39, or 40-64; female; current and past being treated for psychological problems; past treatment for physical diseases; loneliness, frustration, COVID-19-related anxiety, or COVID-19-sleepless. Protective factors were optimism (OR = 0.76, 95% CI = 0.71-0.80, *p* < 0.001) and healthy sleep habits (OR = 0.87, 95% CI = 0.83-0.91, *p* < 0.001).

### Comprehensive interaction structure of psychosocial variables associated with psychological distress

The final convergence results of the non-parametric Bayesian co-clustering are shown in Figure 3. Twenty-two psychological distress clusters were extracted, of which six clusters consisted entirely of those with SPD, four clusters consisted entirely of those with MMPD, and seven clusters consisted entirely of those with NPD. The characteristic interaction structures that influence psychological distress severity are summarised below.

**Figure 3.**
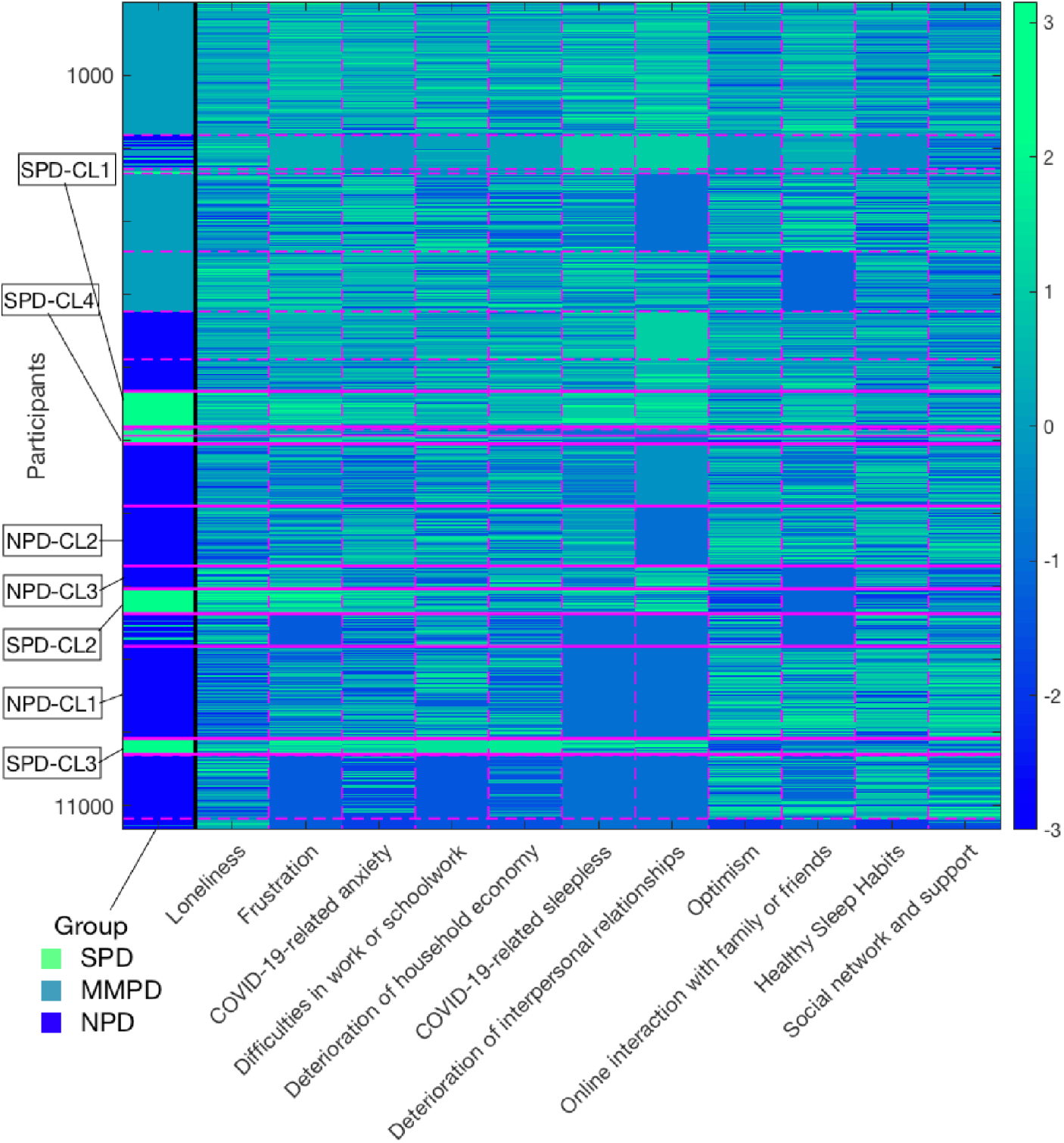
Comprehensive interaction structure of psychosocial variables associated with psychological distress. Rows represent the participants data and columns represent the groups of psychological distress and features about COVID19-related psychosocial factor. The magenta dotted or solid lines indicate the division of each cluster. The color bar indicates the z-score of features.

First, the largest cluster consisting of the SPD group (SPD-CL1, n = 485, 37.2%) showed particularly severe deterioration of relationships with familiar people and COVID-19-related sleeplessness. The second-largest cluster, consisting of the SPD group (SPD-CL2, n = 341, 26.2%), showed particularly high levels of loneliness and frustration, and a lack of online interaction with family or friends and optimism. The third-largest cluster, consisting of SPDs (SPD-CL3, n = 215, 16.5%), showed highly pronounced difficulties in work or schoolwork and deterioration of household economy. In the fourth-largest cluster composed of SPDs, the high level of COVID-19-related anxiety was pronounced. Frustration was also high in all clusters. Taken together, frustration and the combination of individual factors, such as loneliness and household economy, were associated with more severe psychological distress.

Contrastingly, the largest cluster in the NPD group (NPD-CL1, n = 1,261, 21.4%) had high levels of difficulties in work or schoolwork, while they were also highly optimistic, engaged in extensive online interactions, and maintained healthy sleep habits. In the fourth-largest cluster, composed of the NPD group (NPD-CL2, n = 821, 14.0%), COVID-19-related anxiety and deterioration of household economy were indicated, while high levels of optimism and social networks were characteristic of the cluster. In the NPD-CL3, which consisted of the NPD group (n = 317, 5.4%), there was a pronounced deterioration of relationships with familiar people and a high level of loneliness, while the cluster was characterised by low difficulties in work or schoolwork and low COVID-19-related anxiety. Taken together, even if risk factors for severe psychological illness were present, a low number of risk factors and the presence of protective factors were associated with lower psychological distress.

## Discussion

The purpose of this study was to report the distribution of psychological distress severity ∼1 month after the initiation of mild lockdown by the declaration of a state of emergency (7 April to 12 May 2020), and to elucidate the risk and protective factors for psychological distress associated with mild lockdown.

The study was conducted before the mild lockdown was phased out, and the seven major cities where the mild lockdown was initially implemented were included in the data collection.

Based on K6 and PHQ-9 scores, ∼50% of participants were distressed and ∼20% were depressed. Regarding the distribution of K6 scores, the proportion of those with psychological distress was significantly higher when compared to previous national survey (CSLC) data from 2010, 2013, 2016 and 2019. This suggests that the one-month mild lockdown adversely effected the mental health of the population, consistent with previous studies reporting increases in depression, anxiety, and stress during the lockdown ^2,3,5,6^.

The groups most likely to be significantly affected by mild lockdown were healthcare workers, those who were currently or previously treated for psychological problems and younger participants (aged 18-19 and 20-39 years). In these groups, there was a significantly larger proportion of those with MMPD or SPD and a significantly higher level of psychological distress.

Healthcare workers experienced increased psychological distress, especially MMPD, consistent with previous studies reporting a higher risk to healthcare workers ^10,42^. It is assumed that a variety of factors affect psychological distress among healthcare workers, including demanding work, fear of infection, moral injury ^43^, and stigma ^8^. Communication, adequate rest, and practical and psychological support in the workplace may be protective factors against psychological distress ^11^. Therefore, it is important for institutions to establish a systematic support system for healthcare workers. Additionally, approaches such as providing the public with sufficient reliable information to counter stigma against healthcare workers are necessary.

History of treatment for psychological problems was the most significant risk for psychological distress severity. In China and Italy, exacerbations of psychological distress, such as stress and depressive symptoms, have also been reported in patients with psychiatric disorders during lockdown ^44,45^, suggesting that mild lockdown may lead to exacerbation or recurrence of psychological symptoms. A variety of factors can be attributed to this, including excessive fear of infection, lack of access to healthcare services such as home isolation and the closure of daycare facilities, and increased economic hardship ^44,46^. Therefore, especially for patients previously treated for psychiatric disorders, attending physicians should pay special attention to the psychological health of the patient and provide prophylactic support to prevent exacerbation of psychological distress and psychiatric symptoms. For this purpose, it is important to establish a system that enables remote support, including medical treatment, psychological assessment, psychotherapy, and health guidance, using online and telephone services.

Comparing age groups, younger adults were at particularly high risk, consistent with studies from China ^47^ and Spain ^4^. For these individuals, psychological distress may be influenced by the loss of various academic opportunities; anxiety about schooling, graduation, and future prospects; financial difficulties due to the lack of part-time work; and high exposure to social media. Considering the significantly larger proportion of students in the SPD group, it is important that educational institutions compensate students for their educational opportunities and pay particular attention to their mental health. Additionally, it is important for the government and stakeholders to provide information and other support to students to alleviate concerns about employment. Difficulties in work and frustration contributed to distress severity. Therefore, employers should be proactive in their approach to younger-aged professionals to relieve such barriers and promote their mental health.

In contrast, those ≥65 years had the lowest psychological distress of all age groups. This can be attributed to the fact that the elderly maintained the best lifestyle habits, such as exercise, healthy eating, and sleep, and had the lowest levels of frustration and anxiety about COVID-19 compared to other age groups (Supplementary Table 1). Since Japan has the highest proportion of elderly in their population worldwide ^48^, health education on lifestyle for the elderly is popular in the country, and this education may have played a preventive role. Additionally, many older adults do not have access to social media^49^, and these results may have contributed to reducing the increase in anxiety about COVID-19. Furthermore, because Japanese people have traditionally refrained from acting out in consideration of their surroundings ^50^, the elderly, especially those with such cultural considerations, may be less susceptible to the frustrating effects of mild lockdown. Since previous studies have shown inconsistencies in the effects of lockdown across age groups ^6^, future research should take into account lifestyle, social media usage, and cultural background.

Based on the distribution of psychological distress, other populations that require consideration are women, singles, and low-income individuals. Consistent with this study, previous research has shown that being a woman ^2,4,12^ and single ^47^ are risk factors for poor mental health during lockdown. Among those experiencing quarantine, participants with a relatively low total household income have significantly higher post-traumatic stress and depressive symptoms than their counterparts ^51^. A variety of factors can be assumed to underlie the high risk of these populations (e.g. difficulties in living with single mothers). Therefore, it is important to understand these individuals’ difficulties in detail during mild lockdown, and additional social support, such as help from social workers, should be considered.

Psychological risk factors for psychological distress included COVID-19-related sleeplessness, COVID-19-related anxiety, and frustration. In contrast, this study revealed that healthy sleep habits and high levels of optimism were protective factors against psychological distress. Sleep problems are widely known to affect mental health ^52^, and this study also indicated that sleep can be both a risk and protective factor for psychological distress. Therefore, establishing stable and healthy sleep habits may be important as a preventive approach to psychological distress.

Furthermore, because COVID-19-related sleeplessness is also closely linked to COVID-19-related anxiety ^53^, an approach aimed at anxiety reduction may be useful in improving both sleep and anxiety. Since higher levels of satisfaction with information about COVID-19 are associated with lower levels of anxiety regarding COVID-19^2^, it would be useful to disclose appropriate information to people.

Interestingly, previous studies have not focused on the usefulness of optimism as a protective factor. As an approach to increasing optimism as a protective factor, it is important for governments and social media to first communicate the prospects, based on scientific and objective information. Additionally, governments should implement rapid and extensive support policies for people, businesses, and institutions in difficult situations. Furthermore, cognitive-behavioural therapy may be useful for people with excessive anxiety and pessimism. These approaches are expected to reduce anxiety caused by uncertainty about the future, which is expected to contribute to increased optimism and, consequently, to be a protective factor against psychological distress.

A comprehensive mapping of psychological distress severity and the structure of interactions between psychosocial variables revealed that there are various dynamics of difficulties behind psychological distress. In particular, factors such as high levels of loneliness, deterioration of relationships with familiar people, COVID-19-related sleeplessness, increased COVID-19-related anxiety, deterioration of household economy, and work and academic difficulties characterised the main SPD clusters. Although these risk factors were present, the absence of overlapping risk factors and the presence of protective factors were characteristic of the main NPD clusters. The results suggest the importance of an approach that reduces the variety of psychosocial risk factors faced by each individual and also fosters protective factors.

Given the diverse backgrounds of psychological distress caused by mild lockdown, a collaborative and cross-disciplinary approach by a variety of agencies is crucial to provide optimal support for individuals’ difficulties. In other words, it is essential for federal and local government agencies and institutions in the fields of industry, medicine, welfare, and education to work together in a flexible manner to focus on the difficulties of the individual. For instance, for people with significant loneliness and deterioration of household finances, it is necessary to establish a support system that can alleviate these concerns; for example, strengthening social support in communities, workplaces, and medical institutions as well as guaranteeing wages. To address these diverse difficulties, creating a cross-disciplinary support agencies/online platform that provides easy access to all information regarding support during a mild lockdown can be useful, allowing for rapid provision of support tailored to individuals’ problems.

This study had some limitations. Given that we employed a cross-sectional design, it is difficult to examine the long-term impact of mild lockdown and the causal effect of risk and preventive factors. Longer-term follow-up is needed to clarify the evolution of prevalence and causal relationships, such as what variables mitigate or exacerbate the effects of mild lockdown. Additionally, while the results indicate demographic characteristics that may be risk factors for psychological distress, the analysis remains at an exploratory level, because this study provides a preliminary report of the effects of mild lockdown. Considering that different psychological burdens among healthcare workers depend on their job duties ^54^, further elaboration with specific groups is needed. Considering the stressful situations and problems specific to each group would allow the proposal of strategies optimised for each group to effectively alleviate psychological distress. Moreover, because we obtained data only from an online survey, the psychological distress of those without online access remains unexamined. Therefore, it is necessary to combine other methods besides online research to improve the generalisability of the results. Finally, it is difficult to make a simple comparison of the magnitude of the impact of lockdown with coercion and mild lockdown. To make a detailed comparison, consideration of various differences between studies, such as the human suffering caused by COVID-19, the timing of the survey, and the extent and duration of the lockdown is necessary. Therefore, it is desirable to accumulate further research and to implement an integrated research approach that examines the differences in the effects of lockdown with and without coercion.

Despite these limitations, this study provides ample data from seven major cities where the highest numbers of cases were reported during the implementation of mild lockdown following the declaration of a state of emergency. Given that retrospective studies suffer from the effects of recall bias, the current data—collected during the implementation of the mild lockdown and examined ∼1 month from implementation to just before the lockdown was lifted—may prove useful in clarifying the impact of the mild lockdown. The findings could be used during future periods of infection spread to inform how to help vulnerable populations. Specifically, this study sheds light on how to protect individuals’ mental health during lockdowns and on the effective implementation of various evidence-based policies and approaches.

In conclusion, ∼50% of people in major cities in Japan reported mild or greater psychological distress during their one-month mild lockdown experience. This percentage was larger than that observed in previous national surveys. The effects were particularly pronounced among healthcare workers, those with a history of treatment for psychiatric disorders, and younger adults. It was also indicated that support should be considered for women, students, singles, and low-income individuals. Among the psychological variables, COVID-19-related sleeplessness, COVID-19-related anxiety, and frustration were risk factors for increased psychological distress, while healthy sleep habits and optimism were protective factors. Mapping the structure of the interaction of psychosocial variables revealed that there were various backgrounds of psychological distress, indicating the need for specific intervention strategies tailored to each individual’s problem structure. The results suggest that cross-disciplinary public-private sector efforts are important to address individuals’ mental health issues arising from lockdown.

## Data Availability

The raw data supporting the conclusions of this manuscript will be made available by the authors, without undue reservation, to any qualified researcher.

## Acknowledgements

This work was supported by JSPS KAKENHI (grant number 18K13323 and 20K10883); and Project for Creative Research of the Faculty of Integrated Science, Tokushima University. The funders had no role in study design, data collection and analysis, decision to publish or preparation of the manuscript.

## Authors’ contributions

Conceived and designed the study: TY CU NS. Performed the study: TY NS. Analyzed the data: TY NS. Wrote the paper, contributed to and have approved the final manuscript: TY CU NS JY EM.

---

**Supplementary Table 1.** Psychometric characteristics by age group.

| Psychometric data, *M (SD)* | Total | Age group |  |  |  | *F* | df | *p* |
| --- | --- | --- | --- | --- | --- | --- | --- | --- |
|  |  | 18-19 | 20-39 | 40-64 | ≥65 |  |  |  |
| Psychological distress, loneliness and social network |  |  |  |  |  |  |  |  |
| K6 | 5.58 (5.43) | 6.87<sup>a</sup> (5.97) | 6.67<sup>a</sup> (5.76) | 5.46<sup>a</sup> (5.35) | 3.09 (3.65) | 159.22 | (3, 11329) | <0.001 |
| PHQ-9 | 4.90 (5.53) | 6.44<sup>a</sup> (6.25) | 5.95<sup>a</sup> (5.79) | 4.79<sup>a</sup> (5.53) | 2.45 (3.66) | 147.82 | (3, 11329) | <0.001 |
| UCLA-LS3 | 23.5 (5.70) | 23.70<sup>a</sup> (5.54) | 23.54<sup>a</sup> (5.70) | 23.94<sup>a</sup> (5.66) | 21.18 (5.32) | 92.81 | (3, 11329) | <0.001 |
| LSNS-6 | 10.6 (6.17) | 13.13<sup>a</sup> (6.48) | 11.31 (6.01) | 9.83<sup>a</sup> (6.11) | 11.39 (6.38) | 64.33 | (3, 11329) | <0.001 |
| Lifestyle and stress management during mild lockdown |  |  |  |  |  |  |  |  |
| Exercise | 4.17 (1.81) | 4.26 (1.92) | 4.21<sup>a</sup> (1.79) | 4.04<sup>a</sup> (1.82) | 4.59 (1.76) | 37.68 | (3, 11329) | <0.001 |
| Healthy eating habits | 4.34 (1.56) | 4.01<sup>a</sup> (1.68) | 4.25<sup>a</sup> (1.57) | 4.28<sup>a</sup> (1.55) | 4.82 (1.43) | 54.95 | (3, 11329) | <0.001 |
| Healthy sleep habits | 4.63 (1.79) | 3.82<sup>a</sup> (2.00) | 4.24<sup>a</sup> (1.84) | 4.70<sup>a</sup> (1.74) | 5.40 (1.54) | 165.38 | (3, 11329) | <0.001 |
| Activity | 4.03 (1.67) | 4.73 (1.73) | 4.10<sup>a</sup> (1.70) | 3.88<sup>a</sup> (1.66) | 4.38 (1.59) | 46.77 | (3, 11329) | <0.001 |
| Offline interaction with family or friends | 3.53 (1.88) | 4.17<sup>a</sup> (2.02) | 3.52 (1.95) | 3.50 (1.84) | 3.61 (1.79) | 6.99 | (3, 11329) | <0.001 |
| Online interaction with family or friends | 3.27 (2.00) | 4.34<sup>a</sup> (2.14) | 3.85<sup>a</sup> (2.05) | 2.91<sup>a</sup> (1.88) | 3.17 (1.98) | 193.29 | (3, 11329) | <0.001 |
| Altruistically motivated preventive behaviours of COVID-19 | 5.58 (1.66) | 5.87 (1.45) | 5.72<sup>a</sup> (1.60) | 5.48 (1.70) | 5.57 (1.67) | 16.84 | (3, 11329) | <0.001 |
| Optimism | 4.06 (1.57) | 4.26 (1.74) | 4.08<sup>a</sup> (1.65) | 3.94<sup>a</sup> (1.54) | 4.45 (1.39) | 41.27 | (3, 11329) | <0.001 |
| Stressors related to mild lockdown |  |  |  |  |  |  |  |  |
| Deterioration of household economy | 3.80 (1.83) | 3.95<sup>a</sup> (1.93) | 3.92<sup>a</sup> (1.89) | 3.85<sup>a</sup> (1.80) | 3.24 (1.66) | 51.41 | (3, 11329) | <0.001 |
| Deterioration of relationships with familiar people | 2.38 (1.54) | 2.45<sup>a</sup> (1.66) | 2.36<sup>a</sup> (1.58) | 2.47<sup>a</sup> (1.55) | 2.00 (1.33) | 36.13 | (3, 11329) | <0.001 |
| Frustration | 3.31 (1.75) | 3.52<sup>a</sup> (1.84) | 3.65<sup>a</sup> (1.80) | 3.28<sup>a</sup> (1.71) | 2.54 (1.54) | 143.69 | (3, 11329) | <0.001 |
| COVID-19-related anxiety | 4.04 (1.70) | 3.98<sup>a</sup> (1.75) | 4.26<sup>a</sup> (1.69) | 4.01<sup>a</sup> (1.68) | 3.57 (1.69) | 59.97 | (3, 11329) | <0.001 |
| COVID-19-related sleeplessness | 2.44 (1.54) | 2.17 (1.55) | 2.47<sup>a</sup> (1.61) | 2.51<sup>a</sup> (1.53) | 2.12 (1.32) | 26.68 | (3, 11329) | <0.001 |
| Difficulties due to the lack of daily necessities | 3.63 (1.85) | 3.39<sup>a</sup> (1.92) | 3.84<sup>a</sup> (1.90) | 3.68<sup>a</sup> (1.80) | 2.86 (1.71) | 103.62 | (3, 11329) | <0.001 |
| Difficulties in work or schoolwork | 3.82 (2.05) | 4.94<sup>a</sup> (1.97) | 4.23<sup>a</sup> (2.07) | 3.84<sup>a</sup> (1.97) | 2.56 (1.79) | 260.75 | (3, 11329) | <0.001 |

COVID-19, coronavirus disease 2019; K6, Kessler Psychological Distress Scale-6; PHQ-9, Patient Health Questionnaire-9; UCLA-LS3, UCLA Loneliness Scale (version 3); LSNS-6, Lubben Social Network Scale (abbreviated version)
<sup>a</sup> Significant difference from the aged 65 ≥ years group ( $p < .05$ )

## References

1. WHO. Coronavirus disease (COVID-19) Situation Report – 137. (2020).

2. Wang, C. et al. Immediate psychological responses and associated factors during the initial stage of the 2019 coronavirus disease (COVID-19) epidemic among the general population in China. Int. J. Environ. Res. Public Health 17, 1729 (2020).

3. Tang, W. et al. Prevalence and correlates of PTSD and depressive symptoms one month after the outbreak of the COVID-19 epidemic in a sample of home-quarantined Chinese university students. J. Affect. Disord. 274, 1–7 (2020).

4. Losada-Baltar, A. et al. ‘We’re staying at home’. Association of self-perceptions of aging, personal and family resources and loneliness with psychological distress during the lock-down period of COVID-19. J. Gerontol. B. Psychol. Sci. Soc. Sci. (2020). doi:10.1093/geronb/gbaa048

5. Moccia, L. et al. Affective temperament, attachment style, and the psychological impact of the COVID-19 outbreak: an early report on the Italian general population. Brain. Behav. Immun. (2020). doi:10.1016/j.bbi.2020.04.048

6. Mazza, C. et al. A nationwide survey of psychological distress among italian people during the covid-19 pandemic: Immediate psychological responses and associated factors. Int. J. Environ. Res. Public Health 17, 3165 (2020).

7. NHK. COVID-19 cases in Japan (NHK report). (2020). Available at: https://www3.nhk.or.jp/news/special/coronavirus/data-all/. (Accessed: 26th May 2020)

8. Brooks, S. K. et al. The psychological impact of quarantine and how to reduce it: rapid review of the evidence. Lancet 395, 912–920 (2020).

9. Holmes, E. A. et al. Multidisciplinary research priorities for the COVID-19 pandemic: a call for action for mental health science. The Lancet Psychiatry 7, 547–560 (2020).

10. Chew, N. W. S. et al. A multinational, multicentre study on the psychological outcomes and associated physical symptoms amongst healthcare workers during COVID-19 outbreak. Brain. Behav. Immun. (2020). doi:10.1016/j.bbi.2020.04.049

11. Kisely, S. et al. Occurrence, prevention, and management of the psychological effects of emerging virus outbreaks on healthcare workers: rapid review and meta-analysis. BMJ 369, m1642 (2020).

12. Qiu, J. et al. A nationwide survey of psychological distress among Chinese people in the COVID-19 epidemic: Implications and policy recommendations. Gen. Psychiatry 33, e100213 (2020).

13. Taylor, M. R., Agho, K. E., Stevens, G. J. & Raphael, B. Factors influencing psychological distress during a disease epidemic: Data from Australia’s first outbreak of equine influenza. BMC Public Health 8, 347 (2008).

14. Wang, P., Laskey, K. B., Domeniconi, C. & Jordan, M. I. Nonparametric Bayesian Co-clustering Ensembles. SIAM Int. Conf. Data Min. 331–342 (2011).

15. Ministry of Health Labour and Welfare. Comprehensive Survey of Living Conditions. (2020). Available at: https://www.mhlw.go.jp/english/database/db-hss/cslc-index.html. (Accessed: 26th May 2020)

16. Kessler, R. C. et al. Short screening scales to monitor population prevalences and trends in non-specific psychological distress. Psychol. Med. 32, 959–976 (2002).

17. Furukawa, T. A., Kessler, R. C., Slade, T. & Andrews, G. The performance of the K6 and K10 screening scales for psychological distress in the Australian National Survey of Mental Health and Well-Being. Psychol. Med. 33, 357–362 (2003).

18. Kessler, R. C. et al. Screening for serious mental illness in the general population. Arch. Gen. Psychiatry 60, 184–189 (2003).

19. Veldhuizen, S., Cairney, J., Kurdyak, P. & Streiner, D. L. The sensitivity of the K6 as a screen for any disorder in community mental health surveys: A cautionary note. Can. J. Psychiatry 52, 256–259 (2007).

20. Prochaska, J. J., Sung, H. Y., Max, W., Shi, Y. & Ong, M. Validity study of the K6 scale as a measure of moderate mental distress based on mental health treatment need and utilization. Int. J. Methods Psychiatr. Res. 21, 88–97 (2012).

21. Kessler, R. C. et al. Mild Disorders Should Not Be Eliminated from the DSM-V. Arch. Gen. Psychiatry 60, 1117–1122 (2003).

22. Kessler, R. C. et al. Trends in mental illness and suicidality after Hurricane Katrina. Mol. Psychiatry 13, 374–384 (2008).

23. Muramatsu, K. et al. Performance of the Japanese version of the Patient Health Questionnaire-9 (J-PHQ-9) for depression in primary care. Gen. Hosp. Psychiatry 52, 64–69 (2018).

24. Kroenke, K., Spitzer, R. L. & Williams, J. B. W. The PHQ-9: Validity of a brief depression severity measure. J. Gen. Intern. Med. 16, 606–613 (2001).

25. Siu, A. L. et al. Screening for depression in adults: US preventive services task force recommendation statement. JAMA - J. Am. Med. Assoc. 315, 380–387 (2016).

26. Heinrich, L. M. & Gullone, E. The clinical significance of loneliness: A literature review. Clin. Psychol. Rev. 26, 695–718 (2006).

27. Cacioppo, J. T., Hawkley, L. C. & Thisted, R. A. Perceived social isolation makes me sad: 5-year cross-lagged analyses of loneliness and depressive symptomatology in the chicago health, aging, and social relations study. Psychol. Aging 25, 453–463 (2010).

28. Koizumi, Y. et al. Association between social support and depression status in the elderly: Results of a 1-year community-based prospective cohort study in Japan. Psychiatry Clin. Neurosci. 59, 563–569 (2005).

29. Arimoto, A. & Tadaka, E. Reliability and validity of Japanese versions of the UCLA loneliness scale version 3 for use among mothers with infants and toddlers: A cross-sectional study. BMC Womens. Health 19, 105 (2019).

30. Kurimoto, A. et al. Reliability and validity of the Japanese version of the abbreviated Lubben Social Network Scale. Japanese J. Geriatr. 48, 149–157 (2011).

31. Russell, D. W. UCLA Loneliness Scale (Version 3): Reliability, validity, and factor structure. J. Pers. Assess. 66, 20–40 (1996).

32. Durak, M. & Senol-Durak, E. Psychometric qualities of the ucla loneliness scale-version 3 as applied in a turkish culture. Educ. Gerontol. 36, 988–1007 (2010).

33. Shevlin, M., Murphy, S. & Murphy, J. The Latent Structure of Loneliness: Testing Competing Factor Models of the UCLA Loneliness Scale in a Large Adolescent Sample. Assessment 22, 208–215 (2015).

34. Zarei, S., Memari, A. H., Moshayedi, P. & Shayestehfar, M. Validity and reliability of the UCLA loneliness scale version 3 in Farsi. Educ. Gerontol. 42, 49–57 (2016).

35. Lubben, J. E. Assessing social networks among elderly populations. Fam. Community Heal. 11, 42–52 (1988).

36. Ceria, C. D. et al. The relationship of psychosocial factors to total mortality among older Japanese-American men: The Honolulu Heart Program. J. Am. Geriatr. Soc. 49, 725–731 (2001).

37. Martire, L. M., Schulz, R., Mittelmark, M. B. & Newsom, J. T. Stability and change in older adults’ social contact and social support: The Cardiovascular Health Study. Journals Gerontol. - Ser. B Psychol. Sci. Soc. Sci. 54B, S302–S311 (1999).

38. Okwumabua, J. O., Baker, F. M., Wong, S. P. & Pilgram, B. O. Characteristics of depressive symptoms in elderly urban and rural African Americans. Journals Gerontol. - Ser. A Biol. Sci. Med. Sci. 52, M241–M246 (1997).

39. Field, A. Discovering statistics using IBM SPSS statistics. SAGE Publications Ltd (2018).

40. RStudio Team. RStudio: Integrated Development for R. (2015).

41. Allen, M., Poggiali, D., Whitaker, K., Marshall, T. R. & Kievit, R. A. Raincloud plots: A multi-platform tool for robust data visualization [version 1; peer review: 2 approved]. Wellcome Open Res. 4:63 (2019). doi:10.12688/wellcomeopenres.15191.1

42. Nickell, L. A. et al. Psychosocial effects of SARS on hospital staff: Survey of a large tertiary care institution. CMAJ 170, 793–798 (2004).

43. Greenberg, N., Docherty, M., Gnanapragasam, S. & Wessely, S. Managing mental health challenges faced by healthcare workers during covid-19 pandemic. BMJ 368, m1211 (2020).

44. Hao, F. et al. Do psychiatric patients experience more psychiatric symptoms during COVID-19 pandemic and lockdown? A case-control study with service and research implications for immunopsychiatry. Brain. Behav. Immun. 87, 100–106 (2020).

45. Iasevoli, F. et al. Psychological distress in serious mental illness patients during the COVID-19 outbreak and one-month mass quarantine in Italy. Psychol. Med. [published online ahead of print, 2020 May 19] (2020). doi:10.1017/S0033291720001841

46. Chaturvedi, S. K. Covid-19, Coronavirus and Mental Health Rehabilitation at Times of Crisis. J. Psychosoc. Rehabil. Ment. Heal. 7, 1–2 (2020).

47. Wang, H. et al. The psychological distress and coping styles in the early stages of the 2019 coronavirus disease (COVID-19) epidemic in the general mainland Chinese population: A web-based survey. PLoS One 15, e0233410 (2020).

48. United Nations Department of Economic Social Affairs Population Division. World Population Prospects 2019: Data Booklet (ST/ESA/SER.A/424). (2019).

49. Ministry of Internal Affairs and Communications. Report on Information and Communication Media Usage Time and Information Behavior in 2018. (2019).

50. Ministry of Health Labour and Welfare. Report on the 2011 Public Opinion Survey. (2012).

51. Hawryluck, L. et al. SARS control and psychological effects of quarantine, Toronto, Canada. Emerg. Infect. Dis. 10, 1206–1212 (2004).

52. Alvaro, P. K., Roberts, R. M. & Harris, J. K. A Systematic Review Assessing Bidirectionality between Sleep Disturbances, Anxiety, and Depression. Sleep 36, 1059–1068 (2013).

53. Ahorsu, D. K. et al. The Fear of COVID-19 Scale: Development and Initial Validation. Int. J. Ment. Health Addict. [published online ahead of print, 2020 Mar 27] (2020). doi:10.1007/s11469-020-00270-8

54. Lai, J. et al. Factors Associated With Mental Health Outcomes Among Health Care Workers Exposed to Coronavirus Disease 2019. JAMA Netw. open 3, e203976 (2020).

